# DRB1 Subtyping Reveals Divergent Risk and Protection for Type 1 Diabetes in Middle Eastern Populations

**DOI:** 10.64898/2025.12.01.25341336

**Authors:** Ikhlak Ahmed, William Hagopian, Muna Sunni, Hajar Dauleh, Hoda Azzam, Hamda Ali, Mohammad Bashir, Khalid Baggar, Dabia Mohanadi, Ajaz Bhat, Jyothi Lakshmi, Shaima Salim, Evonne Chin-Smith, Sura Ahmed Hussain, Michael Weedon, Seth Sharp, Khalid Fakhro, Richard Oram, Ammira Akil

## Abstract

**Background:** Type 1 diabetes (T1D) is strongly influenced by HLA variation, yet current genetic risk models developed largely in European cohorts perform suboptimally in Middle Eastern populations due to region-specific allele frequencies, DR4 subtype heterogeneity, and distinct haplotype structures. We aimed to characterize HLA diversity in the Qatar Biobank (QBB) cohort and develop a Middle East optimized, machine learning–based T1D risk model (MENA T1D-GRS).

**Methods:** We analyzed high-coverage whole-genome sequencing data from >14,000 individuals comprising 7,359 healthy controls, and 410 clinically diagnosed T1D patients plus 230 first-degree relatives (FDR). High-resolution HLA typing was performed using HLA-LA, HLA-HD, and Kourami. Haplotype phasing, LD estimation, and association testing identified population-specific risk and protective configurations. We computed GRS2 using 66 genomic variants and trained an XGBoost classifier integrating 79 weighted HLA features and GRS2 components. Synthetic data augmentation (ADASYN) was applied to correct the class imbalance between T1D cases and controls, thereby enhancing model sensitivity. Model discrimination was evaluated by AUCROC.

**Results:** The QBB cohort exhibited exceptional HLA diversity, with 305 DRB1-DQA1-DQB1 haplotypes. Established risk haplotypes, DR3-DQ2.5 and DR4-DQ8.1 were significantly enriched in T1D cases, with compound heterozygosity conferring >12-fold increased odds. Importantly, DRB1*04:03 was protective (OR=0.54), contrasting sharply with DRB1*04:02 and *04:05. GRS2 achieved an AUC of 0.74 vs. population controls and 0.65 vs. FDRs; AUC improved to 0.81 in autoantibody-positive cases. The MENA T1D-GRS model achieved AUC 0.79 (baseline) and 0.82 with ADASYN. Sensitivity improved to 75–80% in autoantibody-positive subgroups. SHAP analysis revealed allele-specific effects, highlighting the opposing roles of DR4 subtypes.

**Conclusion:** The MENA T1D-GRS provides a population-tailored genomic risk prediction tool, outperforming existing scores and capturing non-linear HLA interactions. It supports early screening, differential diagnosis, and precision medicine efforts in Middle Eastern populations.

## Introduction

T1D is an autoimmune condition driven by complex genetic and environmental interactions. Approximately half of its heritable risk arises from variation within the HLA class II region, particularly DRB1, DQA1, and DQB1 (Redondo et al. 2022). Across global populations, DR3-DQ2.5 and DR4-DQ8.1 haplotypes remain the strongest known genetic susceptibility factors. However, the frequency, structure, and linkage patterns of HLA haplotypes vary widely across ancestral groups (Lenz 2011; Solberg et al. 2008). Consequently, genetic risk prediction models developed in European cohorts often fail to generalize to Middle Eastern and North African (MENA) populations, where unique HLA configurations—including DR4 subtype diversity—alter disease penetrance.

Populations of the Arabian Peninsula show a complex demographic history shaped by admixture, endogamy, and founder effects (Almarri et al. 2021). Studies from Qatar have shown extensive genetic diversity and distinct haplotype structures compared to European populations (Razali et al. 2021). Yet large-scale, high-resolution characterization of T1D-associated HLA features in this region is lacking. Moreover, existing T1D genetic risk scores, collapse DR4 subtypes despite their divergent roles: DRB1*04:02 and *04:05 increased T1D risk, while DRB1*04:03 is protective. This creates a critical need for ancestry-tailored T1D risk models.

This study integrates whole-genome sequencing, high-resolution HLA typing, haplotype phasing, and machine learning to build the first Middle East–optimized T1D-GRS. We leveraged >14,000 QBB participants and a clinically recruited T1D cohort to (1) characterize HLA structure, (2) identify population-specific risk/protective haplotypes, (3) evaluate GRS2 performance, and (4) develop a machine learning model combining HLA features with GRS2 to improve T1D risk prediction in the Middle East.

## Materials and Methods

Study cohorts included two primary datasets: (i) the Qatar Genome Program (QGP) cohort derived from the QBB, and (ii) a clinically recruited cohort of individuals with type 1 diabetes and their first-degree relatives. WGS data were processed using standard GRCh38 pipelines. High-resolution HLA typing was performed using HLA-LA(Dilthey et al. 2019), HLA-HD(Kawaguchi et al. 2017), and Kourami (Lee and Kingsford 2018), with >95% concordance.

Allele frequencies, and haplotype phasing were computed using custom scripts, and haplo.stats package in R (JP and DJ 2023). Association analyses used Fisher’s tests and univariate logistic regression were conducted in R using DescTools package glm() function in base R. GRS2 was computed using the t1dgrs2 Python package available via Bioconda (https://anaconda.org/bioconda/t1dgrs2) with 66 variants (five proxy variants substituted).

We constructed 79 HLA-derived features (alleles, haplotypes, diplotypes), weighted by logistic regression β-coefficients. These were combined with GRS2 components (SCORE, DQSCORE, non-HLA score) and sex. An XGBoost classifier (Chen and Guestrin 2016) was trained using PyCaret (Ali 2021) with 70/30 train–test splits. To address class imbalance, ADASYN synthetic minority oversampling generated ∼4,800 synthetic T1D cases were included in the training set.

Interpretability was assessed via SHAP values, highlighting allele-specific and haplotype-level contributions.

## Results

### HLA Allelic Diversity and Haplotype Structure in the QBB

The QBB cohort exhibited exceptionally high HLA diversity across all classical loci (HLA-A, - B, -C, -DRB1, -DQA1, -DQB1, -DPB1, -DPA1), reflecting the complex population history of the Middle East. The allele frequency spectrum (**Figure 1**) revealed a small number of common alleles (>5% frequency) with a moderate number of intermediate frequency alleles. HLA-B demonstrated the greatest polymorphism among class I genes, with 168 distinct alleles identified but only four exceeding 5% frequency. HLA-A and HLA-C also displayed substantial allelic richness (153 and 96 alleles, respectively), each composed of a limited set of frequent alleles and numerous low-frequency variants. Among class II loci, HLA-DRB1 was the most diverse, with 117 alleles detected, although only six were common. In contrast, DQA1 had a markedly constrained allelic profile, with only six alleles observed at two-digit resolution, indicative of a narrower haplotypic landscape at this locus. Non-classical loci displayed restricted variation; for example, HLA-F was nearly monomorphic (F*01:01 >99%), while HLA-E and HLA-G were dominated by only one or two alleles.

**Figure 1.**
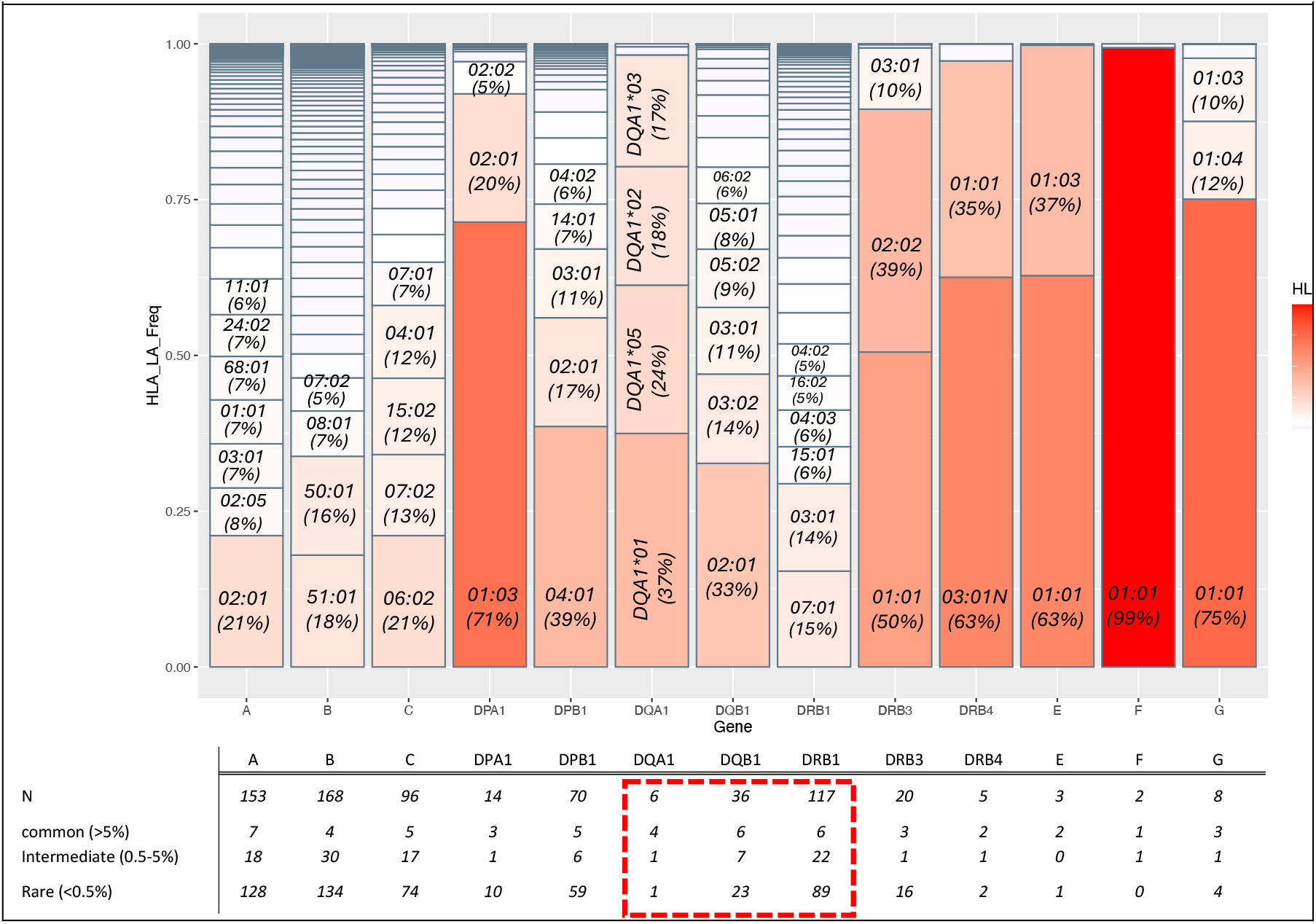
Distribution of HLA allele frequencies across HLA genes in 14,392 individuals from the QBB cohort. Bar plots show allele frequencies across classical and non-classical HLA loci, with shading indicating relative frequency. Top alleles per gene are labeled, and the table below summarizes the number of common (>5%), intermediate (0.5–5%), and rare (<0.5%) alleles. DRB1 shows the highest diversity, while DQA1 is limited to a few dominant alleles. HLA-F is nearly monomorphic. These patterns reflect varying selective and demographic influences across loci.

These patterns are consistent with previously described HLA structures in Middle Eastern and admixed populations and reinforce the unique genetic landscape of Qatar. Several alleles common in the QBB cohort, including HLA-B*51:01, DRB1*03:01, and DQB1*02:01 are also frequent across North Africa and the Arabian Peninsula (Hajjej et al. 2018).

Haplotype phasing identified 305 distinct DRB1–DQA1–DQB1 haplotypes, although the majority were rare (<1% frequency). Only five haplotypes exceeded a 5% frequency threshold, collectively accounting for more than half of all observed haplotypes. These included the well-established autoimmune-associated DRB1*03:01–DQA1*05–DQB1*02:01 (DR3-DQ2.5; 13.8%) and DRB1*07:01–DQA1*02–DQB1*02:01 (13.7%). Other prevalent haplotypes such as DRB1*04:03-DQA1*03-DQB1*03:02 and DRB1*16:02-DQA1*01-DQB1*05:02 were observed at moderate frequencies (5–6%). For the DQA1–DQB1 block, 66 haplotypes were reconstructed, with seven exceeding 5% frequency, highlighting a more constrained haplotypic architecture compared to the three-gene DR–DQ block.

Comparison with global HLA frequency databases highlighted both shared and region-specific haplotype patterns (**Figure 2**). DR3-DQ2.5 frequencies in QBB were similar to those in Mediterranean and North African populations but substantially higher than in East Asian or Sub-Saharan African groups. The DRB1*04:02- and DRB1*04:03-containing haplotypes were moderately common in QBB (∼5%) and nearby MENA populations but significantly more frequent in Ashkenazi Jewish and certain North African groups. These data underscore the distinct immunogenetic landscape of Qatar and provide essential context for interpreting disease association results.

**Figure 2.**
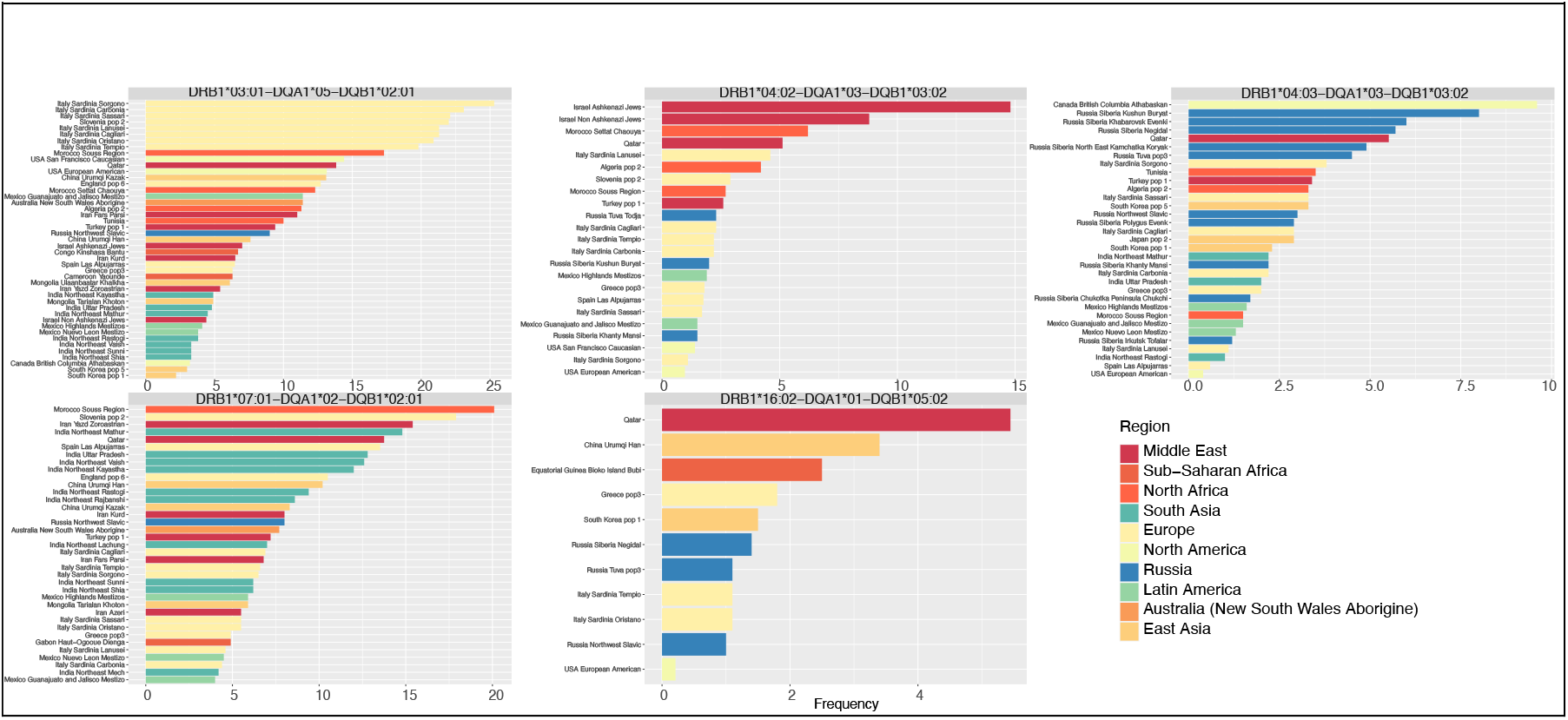
Comparative Frequencies of Common DR-DQ Haplotypes in Global Populations. Bar plots compare the frequency of the top five haplotypes to other global populations using data from Allele Frequency Net Database. Frequencies are shown across major world regions, highlighting both shared and population-specific patterns of haplotype distribution.

**Figure 3:**
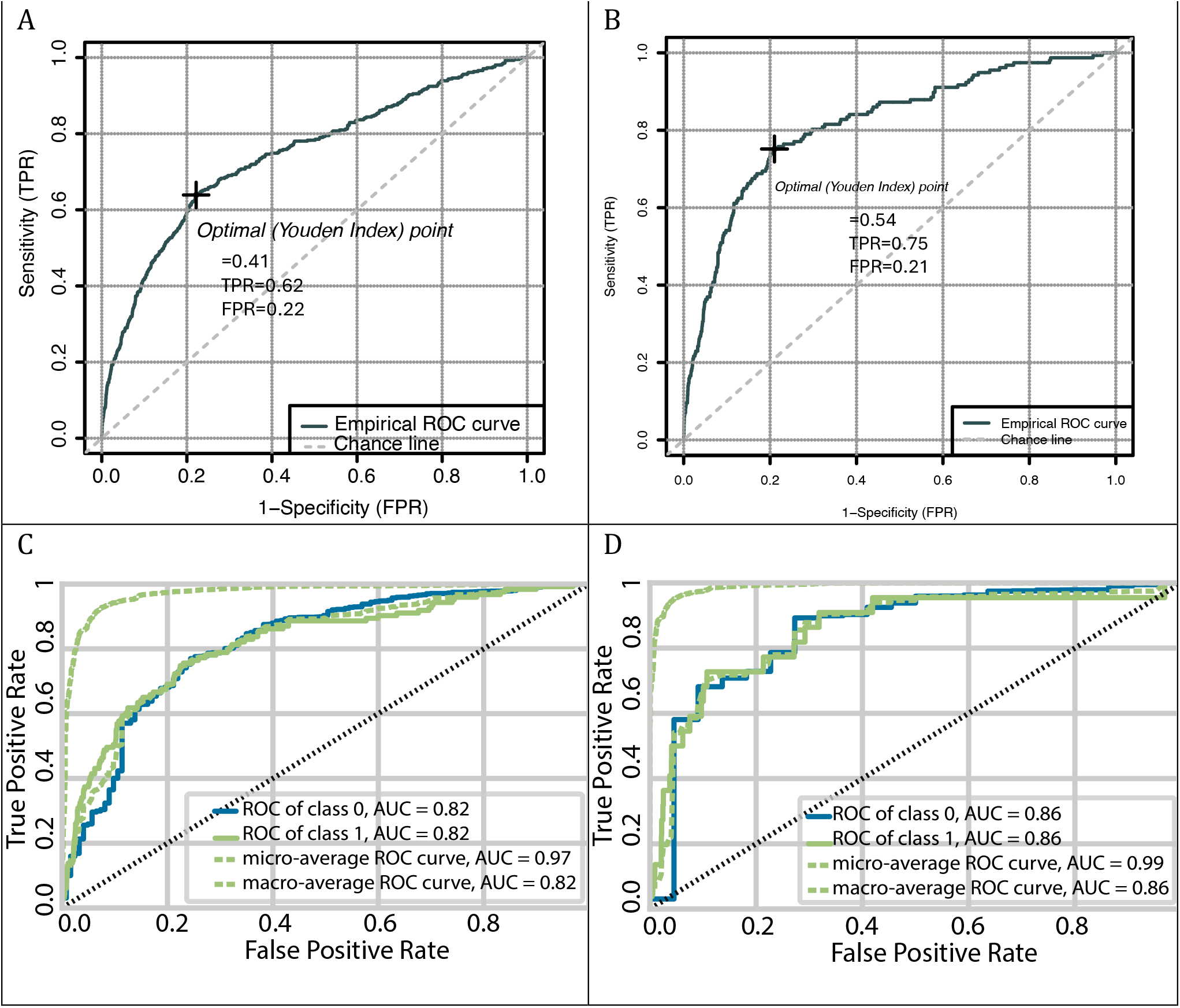
Discriminative performance of GRS2 and MENA T1D-GRS. The top panel shows AUCROC curves evaluating the performance of GRS2 in classifying T1D cases. (A) Compares all cases with population controls (AUC = 0.74), and (B) shows performance restricted to autoantibody positive cases (AUC = 0.81). The bottom panel shows the performance of the XGBoost based MENA T1D-GRS model. (C) presents discrimination for all T1D (class 1) against healthy controls (class 0) in the held-out test set (AUC = 0.82), and (D) shows performance in autoantibody positive cases (AUC = 0.86).

### HLA Association with T1D

We examined DR and DQ haplotypes, diplotypes and serotype combinations in 410 individuals with type 1 diabetes and 7,359 controls to identify HLA features associated with disease risk. Associations were evaluated using Fisher’s test and logistic regression to quantify the effects of both common and low frequency haplotypes.

The strongest risk signal was the DRB1*03:01-DQA1*05-DQB1*02:01 haplotype which corresponds to the DR3-DQ2.5 profile. This haplotype was present in 32.6 percent of cases compared with 13.8 percent of controls, with an odds ratio (OR) of 2.36 (p = 2.51×10^−27^) (**Table 1**). Logistic regression confirmed its robust predictive value (Beta = 1.08, p = 1.16×10^− 42^). Tag SNP analysis identified rs3998177 (G allele, r^2^ = 0.98) as strongly linked to this haplotype, showing 99.9% concordance with the HLA-defined genotype. A second major risk haplotype was DRB1*04:02-DQA1*03-DQB1*03:02 which corresponds to DR4-DQ8. This haplotype occurred in 14.4 percent of cases and 5.2 percent of controls with an odds ratio of about 2.77 (p = 1.47×10^-18^). Several other DR4 containing haplotypes including those with DRB1*04:05 and DRB1*04:01 also showed elevated odds ratios in the range of 2.7 to 4.3 supporting their contribution to T1D susceptibility.

**Table 1.** Significant DRB1-DQA1-DQB1 haplotypes associated with T1D based on Logistic regression analysis.

|  |  |  |  |  |  |  |
| --- | --- | --- | --- | --- | --- | --- |
| Haplotype | Estimate | Std.Error | z.value | p.value | CI.Lower | CI.Upper |
| DRB1*08:03-DQA1*01-DQB1*06:01 | 2.88 | 1.42 | 2.04 | 4.16E-02 | -0.35 | 6.12 |
| DRB1*04:15-DQA1*03-DQB1*03:02 | 2.88 | 1.42 | 2.04 | 4.16E-02 | -0.35 | 6.12 |
| DRB1*15:13-DQA1*02-DQB1*03:01 | 2.88 | 1.42 | 2.04 | 4.16E-02 | -0.35 | 6.12 |
| DRB1*08:04-DQA1*04-DQB1*04:02 | 1.94 | 0.53 | 3.66 | 2.49E-04 | 0.80 | 2.92 |
| DRB1*04:04-DQA1*03-DQB1*03:02 | 1.84 | 0.35 | 5.20 | 2.00E-07 | 1.10 | 2.50 |
| DRB1*09:01-DQA1*03-DQB1*02:01 | 1.79 | 0.58 | 3.09 | 2.00E-03 | 0.51 | 2.85 |
| DRB1*04:01-DQA1*03-DQB1*03:02 | 1.47 | 0.30 | 4.87 | 1.11E-06 | 0.84 | 2.03 |
| DRB1*04:05-DQA1*03-DQB1*03:02 | 1.26 | 0.18 | 7.12 | 1.08E-12 | 0.90 | 1.60 |
| DRB1*04:02-DQA1*03-DQB1*03:02 | 1.25 | 0.12 | 10.81 | 3.00E-27 | 1.02 | 1.47 |
| DRB1*03:01-DQA1*05-DQB1*02:01 | 1.08 | 0.08 | 13.69 | 1.16E-42 | 0.93 | 1.24 |
| DRB1*04:05-DQA1*03-DQB1*02:01 | 1.05 | 0.24 | 4.36 | 1.30E-05 | 0.55 | 1.50 |
| DRB1*15:13-DQA1*03-DQB1*03:02 | 0.92 | 0.23 | 3.96 | 7.40E-05 | 0.44 | 1.36 |
| DRB1*11:04-DQA1*05-DQB1*03:01 | -0.66 | 0.29 | -2.27 | 2.33E-02 | -1.29 | -0.14 |
| DRB1*04:03-DQA1*03-DQB1*03:02 | -0.68 | 0.21 | -3.20 | 1.39E-03 | -1.13 | -0.29 |
| DRB1*13:01-DQA1*01-DQB1*06:03 | -0.75 | 0.26 | -2.87 | 4.10E-03 | -1.31 | -0.28 |
| DRB1*07:01-DQA1*02-DQB1*02:01 | -0.84 | 0.14 | -5.87 | 4.34E-09 | -1.14 | -0.57 |
| DRB1*10:01-DQA1*01-DQB1*05:01 | -0.94 | 0.31 | -3.07 | 2.12E-03 | -1.60 | -0.39 |
| DRB1*16:01-DQA1*01-DQB1*05:02 | -0.95 | 0.36 | -2.65 | 8.10E-03 | -1.74 | -0.32 |
| DRB1*15:13-DQA1*02-DQB1*02:01 | -0.96 | 0.42 | -2.30 | 2.16E-02 | -1.89 | -0.23 |
| DRB1*11:01-DQA1*05-DQB1*03:01 | -1.05 | 0.30 | -3.45 | 5.59E-04 | -1.71 | -0.51 |
| DRB1*16:02-DQA1*01-DQB1*05:02 | -1.30 | 0.28 | -4.65 | 3.35E-06 | -1.91 | -0.80 |
| DRB1*15:03-DQA1*01-DQB1*06:02 | -1.50 | 0.71 | -2.11 | 3.49E-02 | -3.30 | -0.36 |
| DRB1*15:01-DQA1*01-DQB1*06:02 | -1.89 | 0.45 | -4.21 | 2.57E-05 | -2.92 | -1.12 |

Several haplotypes were strongly protective. DRB1*07:01-DQA1*02-DQB1*02:01 and DRB1*04:03-DQA1*03-DQB1*03:02 were significantly enriched in controls with odds ratios of OR = 0.51 (p = 1.08×10^-7^) and OR= 0.54 (p = 1.60×10^-3^) respectively. DRB1*16:02-DQA1*01-DQB1*05:02 was present in 5.7 percent of controls but only 1.6 percent of cases with an odds ratio of about 0.28 (p = 3.61×10^-8^). Other protective haplotypes included DRB1*11:04-DQA1*05-DQB1*03:01, DRB1*13:01-DQA1*01-DQB1*06:03, and DRB1*15:01-DQA1*01-DQB1*06:02, with lower odds ratios and narrower confidence intervals, suggesting consistent and robust effects. Analysis of the corresponding two locus DQA1 DQB1 haplotypes confirmed that DQA1*05-DQB1*02:01 (DQ2.5) and DQA1*03-DQB1*03:02 (DQ8) were enriched in cases whereas DQA1*02-DQB1*02:01 and DQA1*01-DQB1*05:02 were enriched in controls.

Diplotype analysis showed marked differences between high risk and protective genotype combinations. The heterozygous configuration that combines DR3-DQ2.5 with a high risk DR4-DQ8.1 haplotype (DRB1*03:01-DQA1*05-DQB1*02:01/DRB1*04:02-DQA1*03-DQB1*03:02) had one of the largest effects with an odds ratio of about 8.6 (p = 1.93×10^−23^). Homozygous DR3-DQ2.5 and other combinations involving risk associated DR4 alleles (DRB1*04:02 or DRB1*04:05) also showed strong effects with odds ratios above 6 (p < 1.04×10^-8^). In contrast, diplotypes combining protective haplotypes such as DRB1*07:01-DQA1*02-DQB1*02:01 with DRB1*16:02-DQA1*01-DQB1*05:02 were underrepresented among cases and showed a borderline protective association (OR = 0.17, 95% CI: 0 to 0.96, p = 0.046). DRB1*04:03 which is a protective DR4 subtype frequently appeared in diplotypes found only in controls, indicating a strong protective effect.

Serotype based analysis estimated from DQ genotypes showed that individuals carrying both DQ2.5 and DQ8.1 had the highest risk with an odds ratio OR = 8.18 (95% CI: 6.3–10.6, p = 2.6 × 10^-45^). This combination conferred greater risk than either serotype in homozygous form, consistent with a synergistic effect. Protective DQ serotypes were associated with significantly lower T1D risk. Individuals carrying both DQ6.2 and DQ7.5 serotypes showed a markedly reduced odds of disease (OR = 0.06, 95% CI: 0–0.36, p = 1.1×10^-5^). In particular, the DQ6.2 allele (encoded by HLA-DQB1*06:02 on the DRB1*15:01 “DR2” haplotype) was strongly protective: individuals with any DQ6.2-containing genotype were far less common among T1D cases than controls, in line with the well-established protective effect of this allele (Noble 2024).

At the DR serotype level, the DR3 with DR4 risk combination had the strongest association with disease with an OR of 11.77 (95% CI: 8.9–15.5, p = 1.6×10^-55^) for T1D. Not all DR4 alleles contributed equally to risk. DR4 risk alleles such as DRB1*04:02 and DRB1*04:05 were strongly associated with disease, while DRB1*04:03 acted as a protective DR4 subtype. DR4(R+) denotes the subset of HLA-DR4 alleles that are risk-associated (for example, HLA-DRB1*04:02 and DRB1*04:05), whereas DR4(R-) refers to protective DR4 variant, DRB1*04:03, which has been shown to be protective against T1D in other populations as well (Noble 2015, 2024). In our data, the presence of a DR4(R-) paired with DR7 showed the strongest protection. This serotype combination did not occur in any cases. The estimated odds ratio approached zero and the 95 percent confidence interval ranged from 0 to 0.46 (p = 0.0009). Furthermore, genotypes that encoded the DR2 serotype (DRB1*15:0x-DQ6.2 haplotype) were among the most protective at the DR level. For instance, individuals with a DR2/DR4(R-) diplotype (one DRB1*15:01-DQ6.2 haplotype and one DRB1*04:03 haplotype) had a substantially lower risk of T1D than those lacking these protective haplotypes. This observation accords with the known strong protection conferred by the DRB1*15:01-DQB1*06:02 haplotype in T1D (Erlich et al. 2008). In summary, HLA-DR stratification showed that the DR3/DR4(R+) combination is the most disease-predisposing, while carriers of DR4 protective alleles and DR2 haplotypes exhibit significantly decreased risk.

Combined DR and DQ diplotype analysis provided the most fine-grained resolution of risk. The DR3-DQ2.5 together with DR4(R+)-DQ8.1 which unites the full DR–DQ haplotypes corresponding to the DR3 and DR4(R+) serotypes discussed above, had an OR of 12.64 (95% CI: 9.4–17, p = 1.2 × 10-50) for T1D and identified individuals at the highest end of genetic susceptibility. Several protective combinations such as protective DR4(R-) with DR7-DQ2.2 or DR2 with DR7-DQ2.2 were observed only in controls.

Together, these findings show that type 1 diabetes risk in this population is strongly influenced by specific DR and DQ haplotypes and thus underscore the importance of high-resolution HLA characterization for accurate modeling of type 1 diabetes risk.

### Genetic Risk Scores and Machine Learning Risk stratification of T1D

We evaluated the performance of the 67 variant GRS2 model (Sharp et al. 2019) in 410 individuals with T1D, 230 unaffected FDRs and 7,359 population matched controls. GRS2 showed good discrimination between cases and unrelated controls with an area under the curve of 0.74. Accuracy declined when unaffected relatives were used as controls with an area under the curve of 0.65, reflecting shared genetic background. Restricting the analysis to autoantibody positive individuals improved performance, reaching an area under the curve of 0.81 against population controls and 0.73 against relatives.

These findings indicate that GRS2 has value for broad population level stratification.

However, the performance of GRS2 was lower than that reported in European cohorts (Sharp et al. 2019; Luckett et al. 2023). This likely reflects incomplete capture of the regional HLA structure in Middle Eastern populations where several HLA subtypes show divergent effects. Distributions of HLA derived components also showed overlap between cases and controls, indicating the need for population specific optimization.

To address this, we developed the MENA T1D-GRS using a machine learning approach that integrates the GRS2 score with HLA allele, haplotype and diplotype information. We trained multiple ML models using the PyCaret library (Ali 2021), and identified XGBoost (Chen and Guestrin 2016) with consistent performance across datasets. The baseline model trained on 5,408 individuals and tested on 2,318 individuals achieved an area under the curve of 0.79. Because of the strong class imbalance, we used a threshold optimization strategy that balanced accuracy across cases and controls. At the optimized probability cutoff of 0.02, the model achieved accuracy of 73.9% for T1D cases, and 70.0% for controls.

To improve detection of true cases, we used ADASYN (He et al. 2008) to generate synthetic minority class instances (T1D) which increased the training dataset to 10,263 individuals and improved class balance. The enhanced model achieved an area under the curve of 0.82 with higher sensitivity of 75.6% in the test set. Performance was strongest in autoantibody confirmed subgroups. In GAD65 positive individuals the area under the curve reached 0.86 and sensitivity exceeded 77 percent.

These results show that integrating population specific HLA information with machine learning provides improved discrimination. The MENA T1D-GRS offers better sensitivity, greater robustness across disease subgroups and stronger applicability for early risk stratification in Middle Eastern populations.

## Discussion

This study provides the first large scale assessment of HLA diversity and T1D genetic risk in a Middle Eastern population. Consistent with previous reports of high HLA heterogeneity in Arab populations (Hajjej et al. 2018), we observed extensive allelic and haplotypic diversity across classical class I and class II loci. More than three hundred DRB1-DQA1-DQB1 haplotypes were identified, highlighting the complexity of regional immunogenetics and the need for population specific analyses.

Our association analyses confirmed that DR3-DQ2.5 and DR4-DQ8.1 remain major contributors to T1D susceptibility, in agreement with studies in other ancestries (Erlich et al. 2008)(Noble 2015). However, the distribution of DR4 subtypes differed markedly from that seen in European cohorts. DRB1*04:02 and DRB1*04:05 showed strong positive associations with T1D, whereas DRB1*04:03 was protective. Diplotype level results showed that individuals carrying both DR3-DQ2.5 and a risk associated DR4-DQ8.1 haplotypes had the highest odds of disease, while combinations involving protective DR4 or DR2 haplotypes were found mainly or exclusively among controls. These findings support the view that subtype specific variation within the DR4 group plays a central role in disease risk in this region (Noble 2024) and should not be collapsed into broad serotype categories.

We evaluated the performance of GRS2, a widely used type 1 diabetes genetic risk score originally developed in European cohorts (Sharp et al. 2019). GRS2 showed moderate discrimination in this population, with improved performance among autoantibody positive cases. However, accuracy remained below values reported in European datasets (Sharp et al. 2019; Luckett et al. 2023), likely due to incomplete capture of population specific HLA structure which warrants the need for ancestry tailored risk models.

To address this limitation we developed the MENA T1D-GRS using a machine learning approach that integrates GRS2 with detailed allele, haplotype and diplotype level features. XGBoost (Chen and Guestrin 2016) provided the most consistent performance. The baseline model achieved an area under the curve of 0.79, and performance improved to 0.82 after synthetic oversampling with ADASYN (He et al. 2008). Discrimination was strongest in autoantibody confirmed subgroups, achieving an area under the curve of 0.86 for GAD65 positive individuals. These results suggest that incorporating detailed HLA structure and population specific patterns can substantially improve risk stratification beyond existing approaches.

In conclusion, our results highlight substantial regional heterogeneity in HLA structure and its contribution to type 1 diabetes susceptibility. Integrating this population specific information through machine learning improves discrimnation performance and provides a foundation for developing precision medicine tools tailored to Middle Eastern populations. Future work will focus on external validation and the integration of additional genomic and clinical features to refine risk assessment strategies.

## Ethics Approval and Consent to Participate

This study is approved by Sidra Medicine IRB committee. Informed consent was obtained from all participants included in the study through QBB Cohort Study QF-QBB-RES-ACC-0075 QBB IRB Approval number is: Full Board-2017-QF-QBB-RES-ACC-0075-0023. Deidentified research conducted under the project E-2020-QF-QBB-RES-ACC-0150-0143. QBB IRB Approval number, E-2017-QF-QBB-RES-ACC-0026-000.

## Data Availability

The QBB data used in this study is accessible upon application through the Qatar Biobank portal (https://www.qatarbiobank.org.qa/), pending institutional review board approval. Additional data produced in the present study are available upon reasonable request to the corresponding author.

